# Developing and piloting a preterm birth registry prioritizing social determinants of health and patient-reported data

**DOI:** 10.1101/2022.03.21.22272683

**Authors:** Layla Joudeh, Jodi Stookey, Johanna Burch, Rachel L. Kaplan, Sylvia Guendelman, Aline Armstrong, Rebecca Jackson

## Abstract

Biomedical factors and social determinants of health (SDH) affect preterm birth (PTB). Given the complexity of PTB and the increasing rates in the United States, public datasets involving multicomponent variables—from biomedical to structural—can motivate novel interventions to address PTB in the US. The primary aim of this study was to develop a PTB registry based on multi-modal data collection tools that prioritize biomedical and SDH data and identify PTB phenotype (i.e. spontaneous labor, preterm premature rupture of membrane, or medically indicated). The secondary aim of this study was to execute a pilot study to assess feasibility. This study aimed to describe recruitment practices, assess data accessibility and concordance, and to provide an example of how the registry can be used to generate hypothesis and analyze data. We designed the registry through a conceptual model provided by the Dahlgren and Whitehead model of SDH using validated questionnaires and literature on PTB. The registry included a survey, interview, and medical and birth certificate abstraction. To pilot the registry, we recruited 92 participants who delivered preterm, were admitted for risk of preterm delivery, or delivered at term at an urban public hospital. Enrollment was most effective in-person and in the postpartum period. Consent to different parts of the registry was similar regardless of when participants were recruited. There was also a range of data concordance depending on the data source and chosen variable. The registry’s PTB phenotype algorithm identified the correct PTB phenotype 100% of the time. The example analysis demonstrated six unique SDH domains. Participants who delivered preterm reported an average of 11 total stressors and 19.7 protective items and 66% had a significant medical or obstetric comorbidity. Results of this study demonstrate that a PTB registry is feasible and could help advance research to prevent PTB.

## INTRODUCTION

Despite medical advancements in the United States, preterm birth (PTB) rates have been steadily increasing since 2014 (1), and non-Hispanic Black individuals have a 50% higher rate of PTB than other racial and ethnic groups (2). PTB is multifactorial and involves social, structural, environmental, genetic, and biomedical contributions (3). The World Health Organization defines social determinants of health (SDH) as “non-medical factors that influence health outcomes: the conditions in which people are born, grow, work, live, and age, and the wider set of forces and systems shaping the conditions of daily life. These forces and systems include economic policies and systems, development agendas, social norms, social policies and political systems (4).” Decreasing the racial disparity in PTB requires focused efforts on understanding SDH (5,6), including housing, safety, racism, childcare, and work; community and individual support systems; and access to and trust in the medical system. Recent literature has pointed to the need to study how medical, genetic, and SDH factors interact to contribute to PTB (6–8). For example, Hong et al. (8) illustrated that in order to decrease the racial disparities in PTB, SDH should be studied in conjunction with genomics data.

One way to effectively study multiple PTB factors simultaneously is through a comprehensive, open-access data registry. Data registries are a unique platform to collect comprehensive data; their large samples offer access to a wide range of clinicians and scientists and can expedite the research process (9–11). However, to our knowledge, there is no central database that focuses on bringing medical, obstetric, and SDH of pregnant people into the research spotlight. Birth registries typically record medical and demographic information but vary on the inclusion and detail of SDH data. For example, the March of Dimes Database for PTB Research focuses on genomic, transcriptomic, immunological, and microbiome data. While databases like these are helpful in designing interventions that could improve birth outcomes, they lack SDH data (12). Electronic health records (EHR) have been shown to be inaccurate for collection of SDH data in comparison with patient report (13–15). A recent study found that incorporating self-reported SDH data into EHRs improved the accuracy of hospital admission risk estimations (16).

PTB phenotype is also important to consider when designing a PTB registry. There are a variety of potential mechanisms and causes of PTB. Accurate determination of PTB phenotype (e.g. preterm prelabor rupture of membranes (PPROM), spontaneous preterm labor (SPTL), or medically indicated) is critical (17,18), and PTB phenotype has been found to be inaccurate in administrative databases (19–22).

Our study aimed to develop and pilot a PTB registry that incorporates biomedical and SDH data. Within the registry, we also aimed to develop an algorithm that identifies PTB phenotype using medical record data. We then used the registry in a pilot study to assess its feasibility. The pilot study aimed to describe recruitment practices, to assess data accessibility and concordance, and to provide an example of how the registry can be used to generate research questions and analyze data. This registry and pilot study can serve as a framework for designing and implementing a patient-centered PTB registry. A feasible, comprehensive registry can lead to robust research at a local and national level that will help reduce PTB rates and disparities.

## METHODS

An interdisciplinary team including epidemiologists, social workers, public health nurses, physicians, and health professional students designed a comprehensive PTB registry. The registry included a survey, interview, and medical record and birth certificate abstraction.

### Designing the registry

We created the four data collection tools—survey, interview, medical record abstraction, and birth certificate abstraction—based on existing research on risks factors for PTB, using validated instruments when possible, and modified them based on input from a preterm birth community advisory board. We designed our instruments using the Dahlgren and Whitehead model of the main social determinants of health, a social-ecological framework that considers health as a function of individual lifestyle factors, social and community networks, and socio-economic, cultural, and environmental conditions (23). Our goals in designing the instruments were to: (1) develop targeted questions that focus on community-level and individual level biomedical and SDH variables, (2) use multiple sources to collect data in order to compare data consistency across the data sources., and (3) include self-reported data. Our instruments focused on biomedical variables and community and individual level SDH. The data collection tools were iteratively revised based on participant and researcher feedback to improve implementation. In the following paragraphs, we will describe each data collection instrument.

The final survey had 73 questions and spanned community-level and individual-level factors (S1 File). Given the survey was designed as a screening tool, to limit the length of the survey, we chose overarching questions from multi-question validated scales that measure common determinants of health, such as housing instability. The community-level factors were housing, work, financial security, food security, discrimination, social services, care quality, and care access. The individual-level factors were health, health-related behaviors, life stressors, trust in health care system and provider, and resilience factors. We identified survey questions and variables using validated questionnaires that included the 2005 LAMBS, 2017 SOLARS, 2012 PRAMS, 2010 LA HOPE, and 2014 MIHA questionnaires as well as the California Health Interview Survey and San Francisco FIMR (24–30). Survey items included both adverse SDH (stressors) (e.g., housing instability) and potentially positive factors (e.g., utilization of community resources).

The interview was composed of open-ended questions about pregnancy experiences to systematically capture qualitative data that might complement quantitative results from the survey and medical record and birth certificate abstractions. The interview transcripts were coded to match the SDH and individual level factor domains in the other data instruments. A previous study showed how qualitative and survey data can be analyzed in conjunction with one another (35).

The main domains of the medical record abstraction were medical and obstetric co-morbidities, risk factors for preterm delivery, labor and delivery care and diagnoses, prenatal care and diagnoses, lab values, mental health, and substance use (S3 File). Because the PTB phenotype was not directly reported in the medical record, the medical record abstraction also included an algorithm that collected data in order to identify the PTB phenotype (Fig. 3 and S3 File). We also included variables to monitor quality of care such as receipt of aspirin, progesterone, and betamethasone. The birth certificate domains included prenatal care and labor and delivery data, newborn baby medical information, and demographic information about the parent(s) (S2 File).

### Piloting the registry

From October 2017 to March 2019, we recruited pregnant or immediately postpartum individuals at a single urban public hospital in three groups: (1) postpartum after a PTB ; (2) during admission for PTL, PPROM, or other preterm medical indications (e.g. preterm hypertensive disorders), or (3) during a labor and delivery triage visit for PTL evaluation (Fig 1). We also recruited a comparison group of participants who were approached after a term birth with no preterm admission or triage visit for PTL (Fig. 1). We enrolled individuals who were at risk for PTB but had yet to deliver preterm to determine feasibility of enrollment to the registry at different points in the hospitalization. The comparison term group was recruited to demonstrate how a case-control study analyzing PTB could utilize both the PTB registry and a control term group.

**Fig 1.**
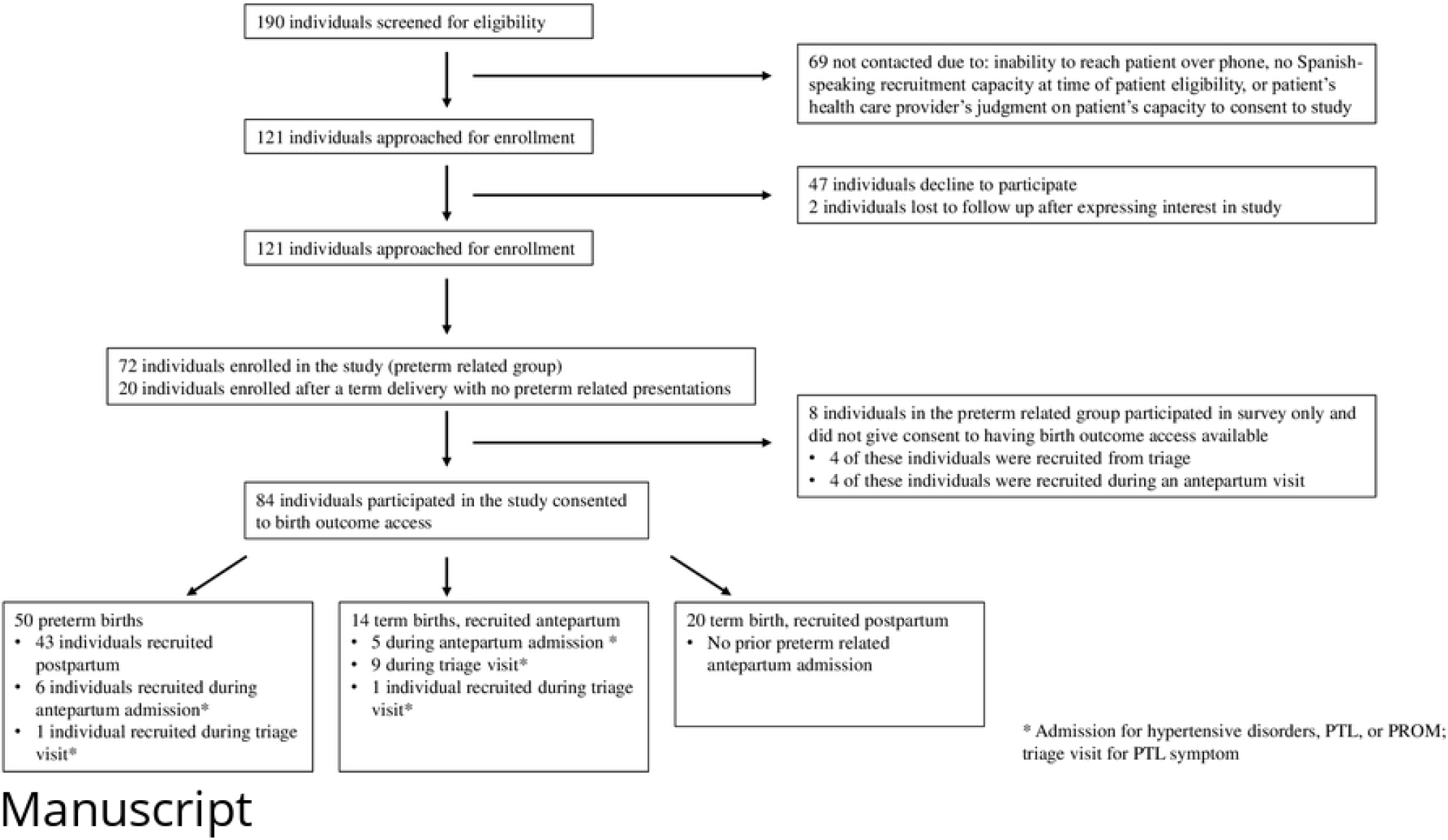
Enrollment and study groups.

Eligible participants were 18 years of age or older, spoke English or Spanish, and had a viable pregnancy beyond 24 weeks gestation. We used convenience sampling and attempted to screen and enroll all pregnant patients who were eligible for the study during our recruiting time period.

### Recruiting for the pilot

We used the EHR to identify eligible participants and contacted them up to three times within one week of the first hospital presentation to share study information. The term group was recruited to achieve similar proportions of race and ethnicity, parity, and language as the preterm groups. Contact was made in person when possible and otherwise via phone calls. To increase autonomy in research procedures, participants were asked to separately consent to each data collection instrument (i.e. survey, interview, medical record abstraction, and birth certificate abstraction). Participants chose when and where to do the survey and/or interview.

Participants completed the survey on paper or on a tablet and had the option to complete the survey alone, with partial assistance, or with a researcher reading every question-and-answer choice. Interviews were completed by phone or in person, recorded, and professionally translated and transcribed. Participants received a $20 gift card for completing the survey and a $30 gift card for completing the interview. Medical record and birth certificate abstractions were completed postpartum to ensure all labor and birth data were available.

### Data management

All study data were stored in Research Electronic Data Capture (REDCap), a secure web platform for building and managing databases. We assessed feasibility with multiple measures. We collected data on enrollment practices (i.e. success rate of enrollment based on if a person was approached in-person versus over the phone; success rate of enrollment based on whether a person was in triage, antepartum, or postpartum). We also recorded time required to complete the survey and medical record abstraction, and asked participants their satisfaction with time to complete the study. We also compared data concordance between birth certificate, medical record abstraction, and survey. We report percent concordance (agreement) and Cohen’s kappa (36,37). Kappa represents the degree of agreement, adjusted for the amount of agreement that could be expected due to chance alone and is commonly interpreted as a high level of agreement (0.81-1.0), substantial (0.61-0.8), moderate (0.41-0.6) and fair/slight (<= 0.40) (36,37).

For purposes of analysis, we characterized risk factors for PTB as major if they were consistently associated with PTB across several studies at an OR above 1.3. These major risk factors include prior preterm birth, multiple gestation, short cervix, hypertensive disorders, renal and cardiovascular disease, diabetes, cholestasis, “Black,” “other race”, BMI <19, age>40 years, tobacco use, and placenta previa or abruption (31–34). We also examined a fuller set of potential risk factors that additionally included substance use disorder, depression, birth spacing <18 months, infection, intrauterine growth restriction, oligo- or polyhydramnios and third trimester bleeding.

To illustrate one possible analysis that can be done using data from the registry, we examined medical and obstetric comorbidities and SDH in three groups: (1) PTB; (2) term births in those who were enrolled during a preterm labor (PTL) triage visit or admission for preterm medical indications, such as PTL, PPROM, or hypertensive disorders; and (3) term births without prior antepartum admission. We categorized SDH variables from the survey, medical record, and birth certificate as potentially protective or as a perceived stressor and grouped them into financial (including housing and food insecurity), employment, health care access and experience, social, health, and personal domains (S1 Appendix). The number of items in each domain ranged from two for employment protective factors to 11 for social stressors (S1 Appendix). Because there were differing numbers of variables in each domain, we normalized on a scale of zero to 10 points for each domain. The average number of stressors and protective factors per person was then calculated for each domain. We evaluated the bivariate association of demographic, medical and SDH with birth group (PTB, term birth after PTL or antepartum admission, term birth with no prior antepartum admission) using Fisher’s exact tests for categorical variables and t-tests for continuous variables. We considered p values less than 0.05 to be statistically significant. We performed all statistical analyses using Stata version 14.1 (38).

## RESULTS

In this section, we first describe the sample characteristics for the registry’s pilot testing. Then we discuss the feasibility of the registry in terms of recruitment, data accessibility and concordance, PTB phenotype. In the final section of the results, we illustrate an example data analysis using data from the registry’s pilot testing.

### Sample characteristics for registry’s pilot testing

We enrolled a total of 92 participants from October 2017 to March 2019 (Fig 1) to pilot test registry. Participants were enrolled prior to delivery (n=29) or after a preterm (n=43) or term delivery (n=20). However, four of the participants recruited in triage and four of the participants recruited antepartum did not consent to sharing their birth outcomes, so their data is excluded from all subsequent analysis. For those enrolled prior to delivery, 11 were enrolled during an antepartum admission for PPROM, PTL or other medical indications and six of the 11 (55%) went on to deliver preterm; 10 were enrolled during a triage visit for PTL, and one delivered preterm (10%) (Fig 1). Of 50 preterm births, 2 (4%) were ≤ 28 weeks gestation, 10 (20%) were 29-33 weeks and 38 (76%) were 34-27 weeks.

### Best practices for recruitment

Recruitment methods were analyzed to determine best practices for an opt-in registry. Enrollment was most effective when participants were approached in person. 55% (n=78) of individuals who were initially approached in person enrolled in the study, while only 21% (n=13) of individuals who were initially approached over the phone enrolled in the study. Comparing antepartum and triage to postpartum enrollment (excluding the comparison term group who were only approached in postpartum), 46% were successfully enrolled in the postpartum period compared with 32% during antepartum admissions and in triage (p=0.04). The average time to complete the survey was 29 minutes (SD=13; range=10-80 minutes), and 94% of survey participants felt it was just the right length. Of participants who completed the survey (n=87), 28% (n=25) asked that the survey be fully read to them and 9% (n=8) had partial assistance. The average time to complete medical record abstraction was 38 minutes (SD=14; range=15-60 minutes).

### Data accessibility and concordance

To facilitate participant autonomy, consent was obtained separately for each part of the registry (i.e. the survey, interview, prenatal records, labor and delivery records, birth certificate data, and the HIPAA-specific topics of mental health, HIV, and substance use). Only 35% consented to all portions of the study. 95% of participants consented to the survey. 82% of participants consented to the interview. 57% of participants consented to obtaining data from the birth certificate. 52% of participants consented to a full medical record abstraction. There were no significant consent pattern differences among those recruited in triage or antepartum, postpartum after a PTB, or postpartum after a term birth. An additional 22% of participants consented to a nearly full medical record abstraction but declined either HIV status, substance use history, and/or mental health information. 26% of participants declined either their complete labor and delivery record and/or prenatal record.

Consistency of information was compared across birth certificate, medical record, and the survey for select variables. Compared to medical record, the birth certificate was found to have >98% agreement and kappa > 0.75 for mode of delivery, chronic hypertension, gestational age at delivery, and induction of labor. Agreement was lower (76-92% concordance and kappa <0.70) for other variables such as prior preterm delivery, PPROM, and use of betamethasone. The birth certificate had both false positives and false negatives. False positives, in which a condition/experience was reported in birth certificate but not confirmed in the medical record, occurred for PPROM (15%) and first prenatal visit ≤8 completed weeks (15%). False negatives, in which the condition was missed in the birth certificate but included in the medical record, occurred for prior PTB (8.5%), first prenatal visit ≤8 completed weeks (15%), diabetes mellitus (2%), PPROM (10%), IOL (3%), use of betamethasone (33%), and hypertension in pregnancy (6%).

There were several differences noted when comparing medical record to survey data. Depression had a very low kappa of 0.14 and a concordance of 67%, with 30% more participants reporting depressive symptoms during their current pregnancy on the survey than carrying a diagnosis of depression in the medical record. On the other hand, 25% of those with a diagnosis of depression in the medical record did not report depressive symptoms during their current pregnancy on the survey. Diagnosis of depression includes both current depression and history of depression. Similarly, unstable housing was reported by 44% in the survey but recorded for only 29% in the medical record (78% concordance, kappa=0.56). Conversely, intimate partner violence was more likely to be reported in the medical record (14%) compared with the survey (7%) (90% concordance, kappa 0.45).

### Preterm Birth Phenotype

Based on comprehensive medical record abstraction, among PTBs, 30% occurred after PPROM, 38% after spontaneous labor, and 32% were medically-indicated (62% of these for hypertensive disorders). Using variables easily accessible in the medical record, we created an algorithm using PPROM (Y/N), induction of labor (Y/N), and Cesarean plus medical conditions known to be associated with indicated preterm delivery (Y/N) (Fig. 3 and S3 File). This algorithm initially misclassified four (8%) of the PTBs labelling them as medically indicated instead of spontaneous labor. The algorithm was revised to include a variable for PTL symptoms (contractions, bleeding) at time of presentation, which was obtained from the history of present illness field and the admission history and physical in the medical record. With this revision, the algorithm correctly categorized all PTBs, as compared with comprehensive medical record review. On the other hand, using only variables available in the birth certificate, 17% of births were misclassified using the algorithm (kappa=0.74).

### Example analysis

A secondary aim of the study was to show how the registry can be used to generate hypotheses and analyze data. In the subsequent section, we show one way to group and describe study participants. Then we present how the registry’s survey, medical record, and birth certificate data can be organized and used for comparative analysis. In our sample pilot, the majority of participants identified as Latinx (57%) or Black/African American (24%), were publicly insured (96%), and had household income <$39,000 (56%) (Table 1). 12% of participants had a prior PTB. 33% of participants had hypertensive disorder(s). 14% of participants had pre-existing or gestational diabetes during the pregnancy (Table 2).

**Table 1:** Selected participant characteristics according to when recruited and delivered, n (%)

|  | **Recruited preterm,**<br>**Preterm birth**<br>**n=50** | **Recruited preterm,**<br>**Term birth**<br>**n=14** | **Recruited**<br>**term, Term birth**<br>**n=20** |
| --- | --- | --- | --- |
| **Age, mean (SD)** | 31.0 (6.5) | 32.4 (4.6) | 31.4 (5.5) |
| **Race/Ethnicity** |  |  |  |
| **Latinx/Hispanic** | 30 (60) | 5 (36) | 13 (65) |
| **Black/African American** | 11 (22) | 6 (43) | 3 (15) |
| **White** | 1 (2) | 2 (14) | 1 (5) |
| **Asian** | 1 (2) | 0 (0) | 1 (5) |
| **Pacific Islander** | 1 (2) | 0 (0) | 0 (0) |
| **Other/Unknown** | 6 (12) | 1 (7.1) | 2 (10) |
| **Study completed in Spanish** | 21 (42) | 3 (21) | 9 (45) |
| **Annual family income** |  |  |  |
| <\$16,000 | 16(48) | 7 (58) | 7 (54) |
| \$16-39,000 | 13(39) | 3 (25) | 1 (7.7) |
| >\$39,000 | 4 (12) | 2 (17) | 5 (39) |
| **Employed during pregnancy** | 29 (62) | 9 (69) | 15 (83) |
| **Housing instability\*** | 13 (27) | 6 (46) | 2 (10) |
| **Food insecurity^** | 17 (36) | 8 (67) | 4 (22) |
| **Did not complete high school** | 10/34 (29) | 3/8 (38) | 4/9 (44) |
| **Needs assistance to complete medical forms** | 9/48 (19) | 1/13(7.7) | 4/18 (22) |
| **Nulliparous** | 15 (31) | 2 (15) | 8 (47) |
| **First prenatal visit by 8 weeks of gestation** | 16/40 (40) | 2/13 (15) | 5/14 (36) |
| **<1 day/week of exercise during pregnancy** | 32 (68) | 3 (23) | 5 (28) |
| **Has someone who can help with daily tasks** | 41 (85) | 10 (77) | 13 (72) |
| **Has someone who can provide emotional support** | 43 (90) | 12 (92) | 16 (89) |

\*Housing insecurity includes houseless or living in shelter, on couch or in SRO; or lives in place without kitchen or private bathroom. ^Food insecurity includes worrying about running out of food or use of food pantry.

**Table 2:** Selected risk factors for preterm birth according to when recruited and delivered, n (%)

|  | **Recruited preterm, Preterm birth n=50** | **Recruited preterm, Term birth n=14** | **Recruited term, term birth n=20** | **p** |
| --- | --- | --- | --- | --- |
| **Prior PTB** | 8 (19) | 2 (14) | 0 (0) | 0.13 |
| **Obstetric/medical comorbidity<sup>^</sup>** | 27 (66) | 10 (71) | 5 (29) | **0.02** |
| **HTN (chronic, gestational, or pre-eclampsia)** | 18 (44) | 5 (36) | 3 (21) | 0.35 |
| **Depression** | 16 (32) | 6 (43) | 8 (40) | 0.69 |
| **Substance use during pregnancy (excluding marijuana)** | 6 (21) | 5 (42) | 2 (14) | 0.28 |
| **Tobacco use during pregnancy** | 5 (12) | 5 (36) | 0 (0) | **0.014** |
| **Black or race defined as “other”** | 17 (34) | 7 (50) | 5 (25) | 0.34 |
| **Any major PTD risk factor\*** | 33 (79) | 10 (71) | 6 (33) | **0.003** |
| **Number of major PTD risk factors\*** |  |  |  | **0.009** |
| • 0 | 9 (21) | 4 (29) | 12 (67) |  |
| • 1 | 12 (29) | 3 (21) | 4 (22) |  |
| • 2-5 | 21 (50) | 7 (50) | 2 (10) |  |
| **Any PTD risk factor<sup>#</sup>** | 40 (91) | 14 (100) | 10 (56) | **0.001** |

<sup>^</sup> Hypertension; diabetes; renal, heart or liver disease; cholestasis; sickle cell disease; seizure disorder; hepatitis C; human immunodeficiency virus (HIV); pyelonephritis; admission for abscess or cellulitis; multiple gestation; placenta previa; short cervix; intrauterine growth restriction; oligohydramnios
\*Major PTD risk factors: risk factors with OR>1.3 in multiple studies, see methods section. Prior PTB, multiple gestation, short cervix, hypertensive disease, Black or other race, age > 40 yo, BMI<19, tobacco use, diabetes, heart or renal disease, placenta previa or abruption.
<sup>#</sup> PTD risk factors include major PTD risk factors plus additional risk factors inconsistently shown in multiple studies: birth spacing <18 months, substance use, depression, asthma, infection (urinary, STI), intrauterine growth restriction (IUGR), oligo/polyhydramnios, third trimester bleeding.

In this section, we illustrate one way to use the registry data to analyze perceived protective factors and stressors that could affect PTB. The frequencies of individual protective factors (n=36) and stressor (n=40) SDH items are shown in S1 Appendix. The most commonly reported stressors among individuals who delivered preterm were: (1) obstetric or medical comorbidities (66%), (2) strenuous work conditions (45.8%), and (3) experiencing poverty as a child (38.3%). Among individual items, the most commonly reported protective factors among individuals who delivered preterm were: (1) during my pregnancy, I had someone I could turn to if I needed someone to comfort me or listen to me (89.6%), (2) during my pregnancy, I had someone to help me with daily tasks (85.4%), and (3) I am satisfied with my prenatal care (81.3%). There were no statistically significant associations found between individual social determinants and delivery group.

The mean number of SDH stressors and protective factors between the three study groups are summarized in Fig. 2. There were no statistically significant differences across delivery groups for each domain or for the total across all domains. Among stressors, the domain with the highest mean score was work (2.84 out of 10). Among protective factors, the highest was health care experience (5.3 out of 10). Participants who delivered preterm reported an average of 11 total SDH stressors and 19.7 total protective factor items of 60 possible (Fig 2). Although not statistically significant, the group that delivered at term but was recruited preterm during admission or triage reported more stressors in most categories than the other two groups.

**Fig 2.**
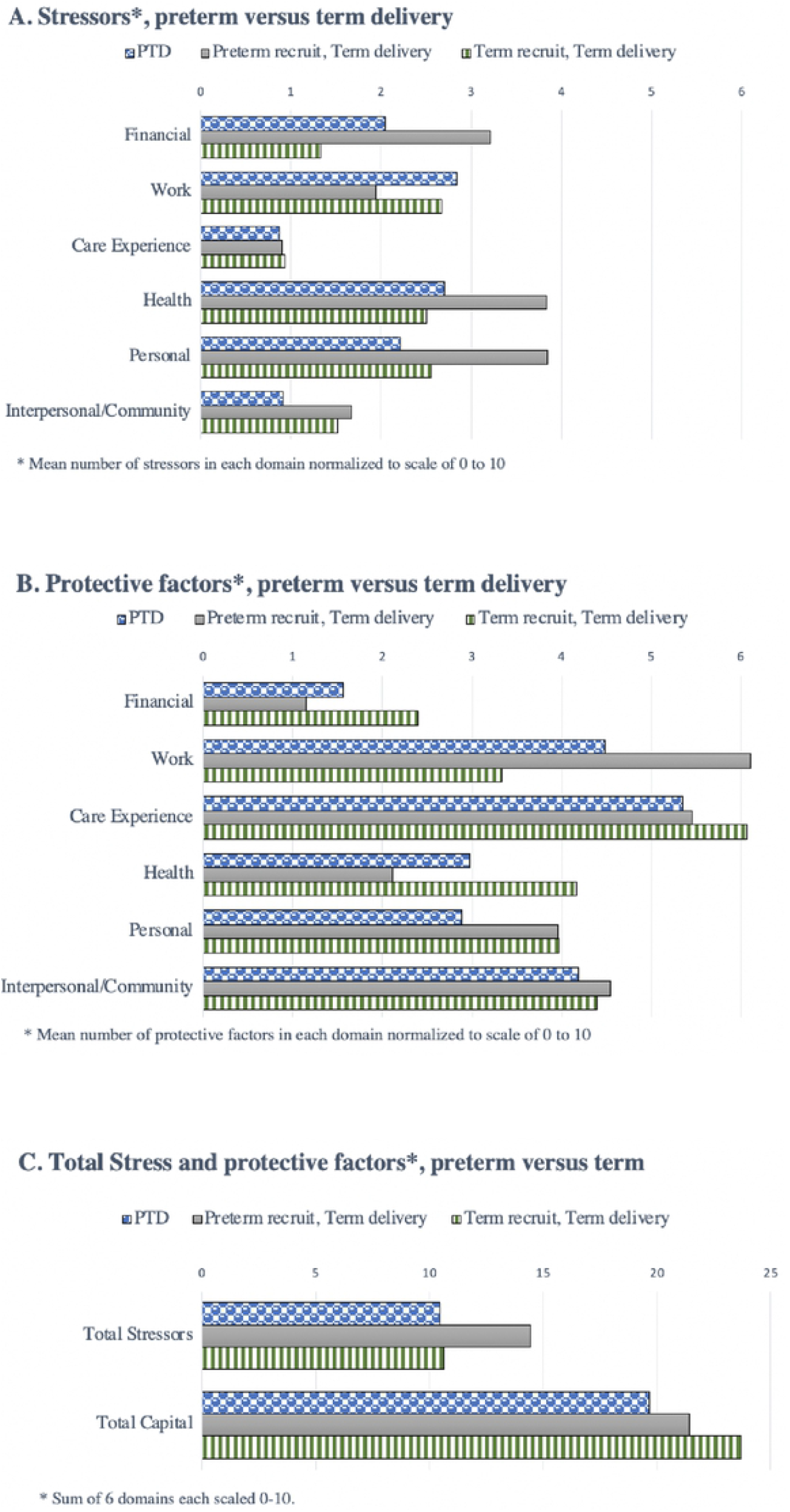
Structural determinants of health distributions

### Individual-level risk factors

Pregnant participants who delivered preterm and those who were recruited preterm but delivered at term had significantly more major preterm delivery risk factors and medical co-morbidities than those recruited at time of a term delivery (p <.05) (Table 2). Among those who had a PTB, 18% had prior PTB, 44% had hypertensive disorder, and 23% had diabetes. There was also significantly more tobacco use during pregnancy (p=.009), obstetrics or medical comorbidity (p=.02), and at least one PTD risk factor (p=.001) in the pregnant people who delivered preterm and those who recruited preterm but delivered at term compared to the participants recruited at time of term delivery.

## DISCUSSION

Our PTB registry pilot highlights methods to develop a feasible PTB registry that captures SDH as well as biomedical data. We also demonstrate how data can be organized and analyzed using a registry. Based on our study, we recommend the following best practices:

### Recruitment practices

For recruiting participants, we recommend in-person, language-concordant recruitment to adequately explain the purpose of the registry, obtain consent, and assist with completion of data collection. One option for public health departments to consider would be to utilize birth certificate clerks to complete registry procedures as they already visit each patient to collect birth certificate data. We also recommend compensating participants for their time and information, as this helps recruitment and engagement and acknowledges the contribution participants have made to medical and public health advancements (39).

While in-person recruitment was effective in our sample, we still encountered varying degrees of consent to specific study procedures. The majority of the participants completed the survey, a data collection tool in which participants can control their responses. In contrast, fewer participants consented to a birth certificate and full medical record abstraction. We were still able to recruit participants using this opt-in process, but studies have shown that opt-out registries yield higher participation (40). The challenge with opt-in and opt-out registries is that varying degrees of consent yields missing data for primary and secondary analysis. However, given mistrust in the medical system, especially in communities of color due to current and historical injustices and discrimination (41–43), it is important to have transparent study procedures such that participants know what data is being collected and how it is used. Thus, a priority for registry design is to have a consensus about the key primary outcome measures that must be collected for all registry participants (10). This will limit data gaps and ensure that when individuals do participate in the registry, their information will contribute to robust multi-site analysis while still facilitating autonomy over their information.

An additional aspect of registry implementation is consensus on what sample to include. Table 2 shows that participants who presented to the hospital with PTB symptoms or conditions that commonly lead to PTB and went on to deliver at term were more similar to those with PTB than with term birth recruited at term. This group included both patients admitted with high-risk conditions (PTL, hypertensive disorders) and patients seen in labor and delivery triage who had PTL symptoms but were not admitted. It was more effective to enroll the admitted patients compared with the triage patients, and there was a higher rate of PTB in the admitted versus the triage patients (40% versus 7%). Including this group in a registry would therefore have the benefit of illuminating the difference between these two groups with different birth outcomes despite having similar symptoms and risk factors for PTB. While including this group would involve more nuanced participant screening, it has the potential of providing more robust data. We also found that participants were more likely to consent to participate if they were approached in the postpartum period, so focusing participant recruitment during this time period could be most effective in optimizing registry participation.

### Data inclusion and analysis

Our results illustrated that self-reported SDH data can be a more reliable way to include SDH data in a registry, as only very limited SDH data is found in the medical record or birth certificate. For the few SDH variables that were available in the medical record, like unstable housing, the medical record under-reported or over-simplified this data. This aligns with previous research that showed that medical records do not accurately capture SDH data (13–15). While medical record abstraction was less accurate for SDH data, it was more accurate than the birth certificate for the majority of medical and obstetric variables, similar to other studies (22,44). Survey and medical record data were complementary in capturing sensitive medical information. Including both medical record and self-reported data in a comprehensive registry would enable researchers to investigate a variety of hypotheses. For example, using our registry data, Reno et al. (35) demonstrated pregnant people receiving behavioral health services had significantly later gestational ages at birth compared to participants in our sample who did not receive behavioral health services.

A registry should also include sufficient data to accurately determine the PTB phenotype, given that PTB is a heterogenous outcome (17,18). While there are alternative options for classifying PTB, the most commonly used is PPROM, SPTL, and other medical indication (17,18,45). Where possible, we recommend updating the EHR to include a drop-down menu that enables streamlined PTB classification. In the absence of classification by the delivery provider, we recommend determining PTB phenotype with EHR data or by using an algorithm (Fig. 3 and S3 File), with inclusion of signs of spontaneous labor (contractions, bleeding) for improved accuracy. We recommend against using birth certificate data to determine PTB phenotype and risk factors associated with PTB. Compared with medical record abstraction, our birth certificate data was misclassified phenotype in 17% and under- or over-reported on a number of other variables. Studies comparing birth certificate data to medical record review similarly show that birth certificate data do not accurately differentiate between PPROM, SPTL, and medically indicated delivery, and under-report medical and obstetric complications (19,46,47).

**Fig 3.**
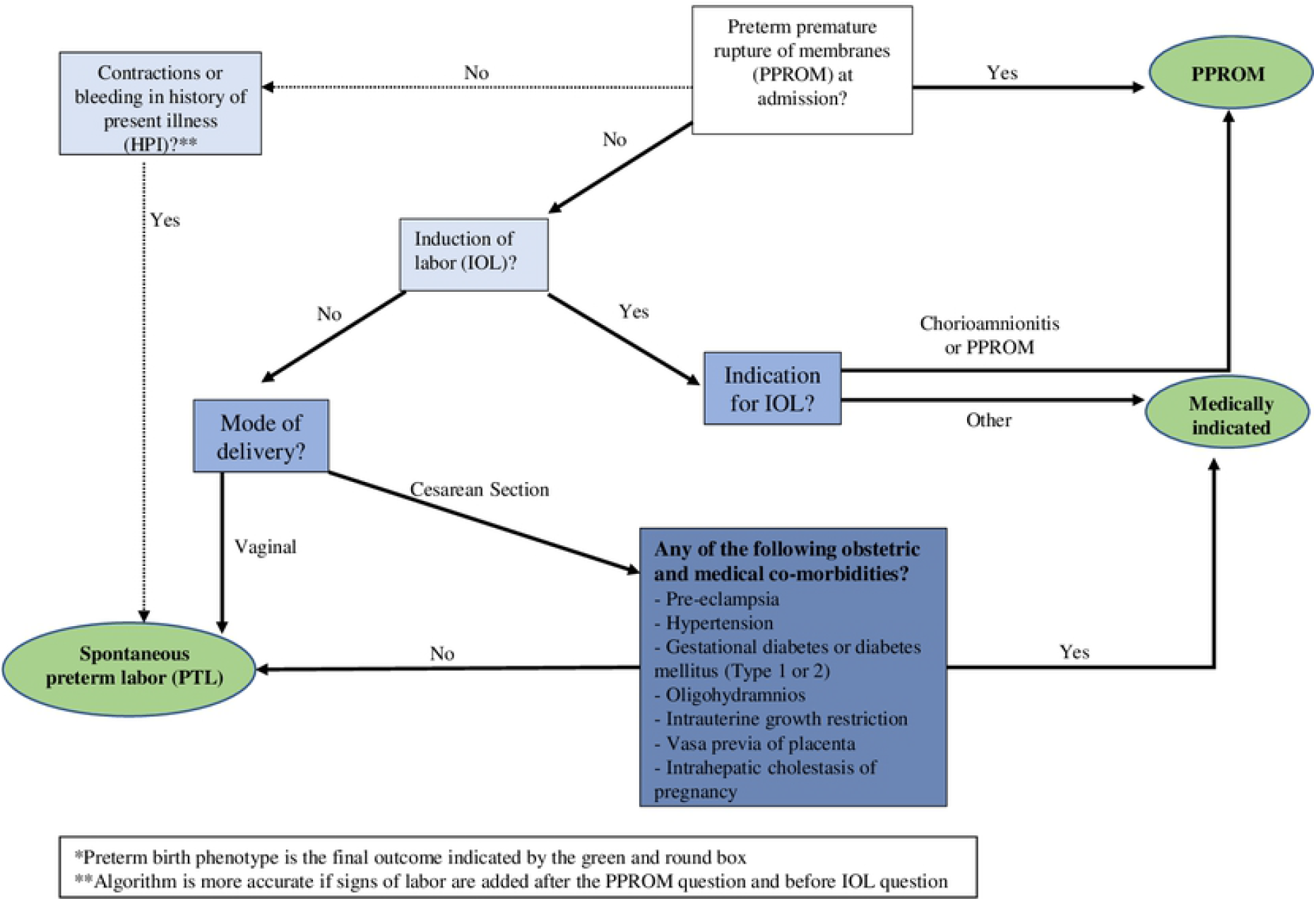
Algorithm predicting preterm birth phenotype*.

Existing studies highlight the need for SDH data in understanding PTB, developing PTB interventions, and improving birth outcomes. Our data analysis illustrates one of many possible analyses using the registry data. A registry comparable to the one described here with large sample size could provide the statistical power necessary to better understand the significant relationships between SDH and preterm birth. Finally, example data analysis demonstrates how a term group could be separately recruited from a PTB registry in order to use the registry to conduct a case-control study. The data could also be adapted for quality improvement purposes. For example, the data could be used to assess and ensure appropriate use of betamethasone and aspirin.

### Future studies

While we have shown that there is a feasible framework for a comprehensive PTB registry, implementing a registry requires logistical support, coordination, and acceptability among health care providers and systems. A future study could assess health care providers’ desires to have a PTB registry and the acceptance of a PTB registry similar to the one presented. Future directions could include using a large sample to conduct robust research that uniquely examines interactions between clinical diagnoses, physical and mental health, SDH, and birth outcomes. Finally, a financial assessment of registry development and maintenance would contribute to the feasibility data provided by these results.

### Limitations

This pilot used a small sample to assess the feasibility. We also did not have the capacity to include participants who spoke languages other than Spanish or English. These limitations raise the possibility of selection bias, as the factors making potential participants unreachable, ineligible, or unwilling to participate could be associated with risk for PTB. The information gathered from these participants and their feedback on how to make the registry more accessible and acceptable are especially important. Lastly, because this study took place at a single site, we cannot generalize the results to other settings. However, despite these limitations, the results described here provide a promising way forward for developing a PTB registry.

## CONCLUSION

A PTB registry incorporating SDH with medical and obstetric factors is feasible. Recruiting participants in person and in the postpartum period, having personnel to support participants in completion of survey, leveraging complementary data sources including self-report and medical records, ensuring transparency in data collection, and providing a participant consent process that promotes individual autonomy will contribute to the development of a patient-centered PTB registry. This patient-centered PTB registry can advance care for pregnant individuals and improve birth outcomes.

## Data Availability

All relevant data are within the manuscript and its Supporting Information files.

## Acknowledgments

We would like to thank our study participants for their invaluable contributions.

## Supporting information captions

S1 Appendix: Domains of social determinants of health and prevalence among study groups

S1 File: Survey data collection instrument in English and Spanish

S2 File: Birth certificate data collection instrument

S3 File: Medical record data collection instrument

